# How detection ranges and usage stops impact digital contact tracing effectiveness for COVID-19

**DOI:** 10.1101/2020.12.08.20246140

**Authors:** Konstantin D. Pandl, Scott Thiebes, Manuel Schmidt-Kraepelin, Ali Sunyaev

## Abstract

To combat the COVID-19 pandemic, many countries have adopted digital contact tracing apps. Various technologies exist to trace contacts that are potentially prone to different types of tracing errors. Here, we study the impact of different proximity detection ranges on the effectiveness and efficiency of digital contact tracing apps. Furthermore, we study a usage stop effect induced by a false positive quarantine. Our results reveal that policy makers should adjust digital contact tracing apps to the behavioral characteristics of a society. Based on this, the proximity detection range should at least cover the range of a disease spread, and be much wider in certain cases. The widely used Bluetooth Low Energy protocol may not necessarily be the most effective option in its current setting.

## Main Text

Caused by the SARS-CoV-2 virus, the Coronavirus disease 2019 (COVID-19) has rapidly spread to a global pandemic with more than 56 Million infections and 1.3 Million deaths worldwide, as of November 2020. Only few countries have managed to minimize the number of infections, and many countries struggle to control the epidemic, some facing a second, even more severe wave of infections *(1)*. A widely accepted means to better control the epidemic is the isolation of infectious individuals. To do this effectively, researchers and governments around the globe have discussed and subsequently introduced digital contact tracing (CT) apps. Once such a CT app is installed on someone’s personal device, it automatically tracks and records information about the individual’s proximity to other individuals that have the same CT app installed on their device. If one individual is diagnosed with COVID-19, this information can be entered into the CT app, which then automatically notifies other individuals that were in physical proximity to the infected person. The notified individuals thus are informed that they have been exposed to the risk of possible infection and may self-quarantine, test, or take other preventive measures to limit the spread of the disease *(2, 3)*.

Many countries have developed and implemented their own CT solutions *(4)*. An important aspect of any CT app implementation is the technique to detect individuals in proximity. Bluetooth Low Energy, for example, is used by many countries *(4)*. However, extant research has opposing views about its suitability for CT. On the one hand, it is among the most accurate technologies for CT *(5)*. On the other hand, lab experiments suggest that a wider distance between smartphones does not necessarily decrease the received signal strength, and Bluetooth Low Energy may register contacts within a relatively wide distance that exceeds the distance in which infections actually happen *(6, 7)*. Beyond Bluetooth Low Energy, extant research on CT has discussed many other technologies to detect the proximity between individuals such as GPS, QR codes *(4)*, or RFID *(8, 9)* (see Table 1). These technologies vary in several characteristics, like their energy consumption, the time interval of physical distance measurement, or the amount of required user action. With regard to epidemic control, the most important differentiating characteristic of these technologies is the proximity detection range (PDR). While some technologies like RFID detect physical contacts only for small distances, the PDR of Bluetooth Low Energy is up to 10 meters *(6)*, and sites-wide QR codes provide snapshots of which persons were at a certain place at a certain time. Given these different characteristics, research has recently called to assess and optimize the suitability of proximity detection technologies for CT apps *(10)*.

**Table 1.**
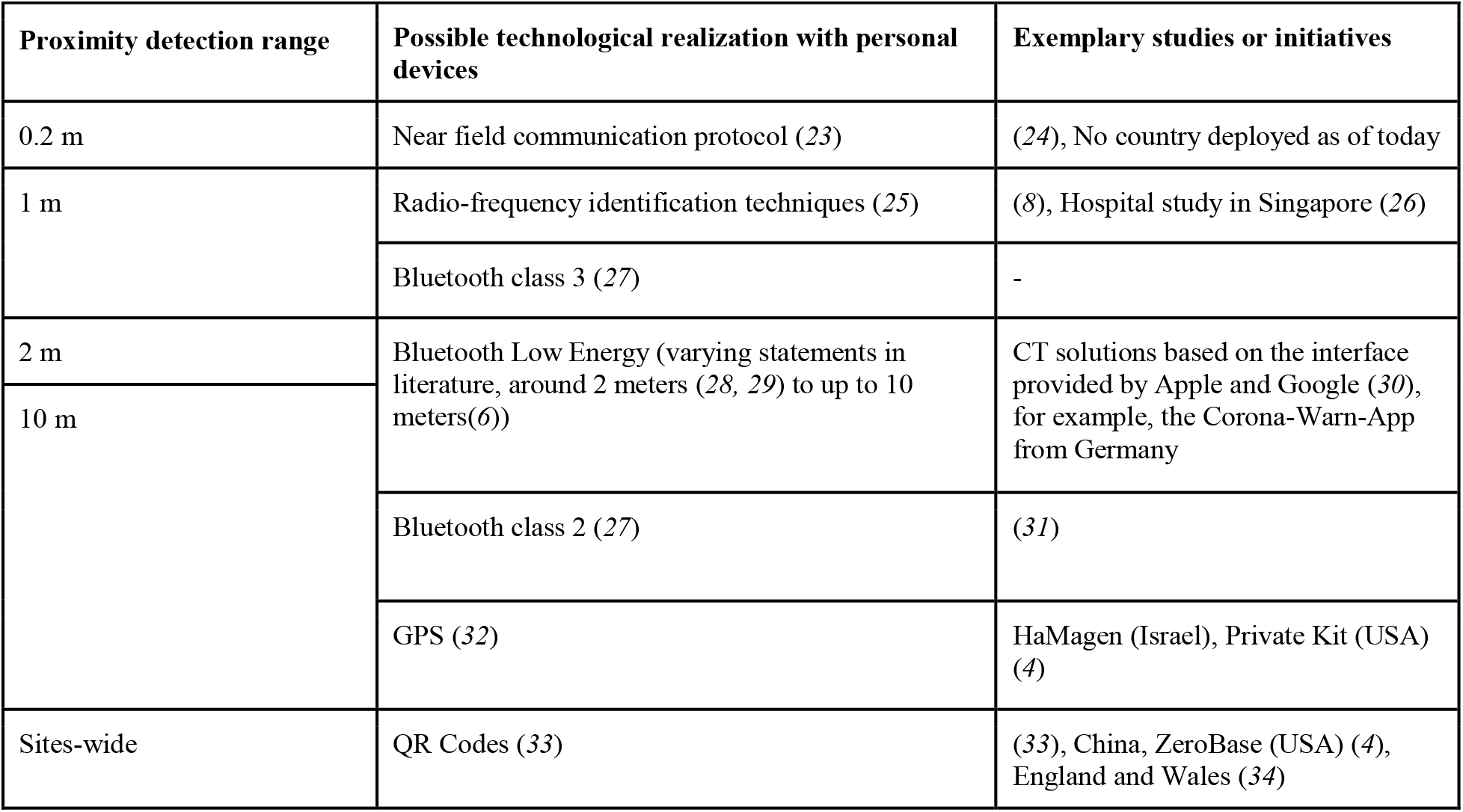
Simulated proximity detection methods and their possible realization with widespread sensors for wearable devices.

In an ideal scenario, CT apps help to detect a contact if an infection event occurred, and do not detect a contact if no infection event occurred. However, in reality no CT technology can achieve this ideal scenario. Instead, the different PDRs of different technologies yield their very own benefits and disadvantages. A PDR wider than a reasonable distance of infection risk may ensure a high recall (i.e., a large share of infectious contacts is registered). However, it may also lead to a low precision (i.e., many of the registered contacts were not actually infected). Such a registration of many contacts may cause large lockdowns. While large lockdowns may be effective to control epidemics, they also yield substantial negative effects such as high economic costs *(2, 11)*, constraints on personal freedom, and harming the populations’ mental health and well-being *(12)*. Furthermore, high shares of false positive contacts that unnecessarily (self-)quarantine may lead to low initial adoption rates of CT apps, and even usage stops, thereby potentially decreasing the overall effectiveness of CT. In contrast, a small PDR may lead to the registration of contacts at risk with a high precision, but it may also miss some relevant contacts, and therefore be less effective in controlling the epidemic. Furthermore, a small PDR enables the localization of individuals more accurately and, thus, potentially opens the door to deducing highly precise movement profiles that impede individuals’ privacy. In some regions, reflections on privacy play a key role in the adoption of CT apps. For example, Norway, has stopped its CT app due to public privacy concerns *(13)*. Given these different considerations, it is not a trivial task for decision makers to decide on the most suitable PDR for CT apps in order to fight the COVID-19 epidemic in their country. Here, we report on a spatial simulation with the aim to analyze how different PDRs implemented in CT apps might influence the course of the COVID-19 epidemic. We thereby also take into account how a usage stop induced by false positive quarantine affects the effectivity of CT apps for different PDRs.

Our work contributes to the scientific knowledge base on COVID-19 and the fight against global pandemics via digital technologies in three ways. First, we evaluate the effectiveness of different PDRs for CT in being able to help bringing the pandemic under control under different initial adoption levels. Second, we study the effect of a usage stop, which may occur once individuals stop using CT due to a false positive quarantine. Third, we demonstrate that spatial simulations can provide valuable insights when studying the effectiveness of CT for combating epidemics.

### Simulation approach

The core of our research method is the spatial simulation of an epidemic where individuals move in a two-dimensional space and thereby can spread the disease. The parameters of the simulated society are set to closely match the German society and are described in detail in the supplementary materials. We chose Germany as a model country for our simulations since it is a country where privacy discussions in general and around CT in particular are very prevalent *(14)*. The severity of the COVID-19 epidemic in Germany falls somewhat medium in international comparison (it does not have the highest infection rates internationally, but COVID-19 is a prevalent health threat there, as of today). Furthermore, Germany has relatively early released a non-mandatory CT app – along with other important measures (e.g., obligations to wear a mask at certain places) – and was able to drastically reduce the number of COVID-19 cases during the summer of 2020. However, as in many other countries, Germany is currently confronted with increasing case numbers (commonly referred to as ‘second wave of infections’) and faces the challenge of how existing measures must be adapted in order to fight the epidemic more effectively. The effectiveness of the German CT app has recently been questioned in the public discourse, due to its number of downloads of ca. 20.3 Million (as of October 2020) *(15)*, resulting in a relatively low estimated adoption rate of ca. 24.5%. Following prior epidemiologic research and evidence where infections are likely to occur *(16)*, we implemented four types of places in order to obtain a mixture of everyday social situations: households, schools, workplaces, and supermarkets. The parameters of our simulation are set such that the base reproduction number R_0_ of the pandemic, without contact tracing measures, has a median of 2.792, which is close to a median value of 2.79 that extant research has identified for the COVID-19 pandemic *(17)*.

The epidemic in the simulation builds on the SEIR model, which states that an individual can be in one of four states: susceptible, exposed, infectious, or recovered *(18)*. Initially, almost all individuals are in an exposed state, and few are exposed to the infectious disease. Exposed individuals then transition into the infectious state after a certain time, in which they can infect other susceptible individuals. After some time, an infectious individual recovers and stays immune to the infectious disease. Prior research showed that the SEIR model is well applicable for COVID-19 *(19)*. At the start of a simulation, ten individuals are exposed, whereas the rest of the population is susceptible. An exposed individual then develops symptoms after the incubation period, which we modeled through a triangular distribution with a mode of 5.5 days, representing a realistic value for COVID-19 *(20)*. However, individuals already transmit COVID-19 around two days before they develop symptoms *(21)*. Accordingly, individuals in the exposed state transition into the infectious state two days before they develop symptoms in our model. Once individuals develop symptoms, we presume that they and their household members go in quarantine for 14 days, the household structure thereby is similar to the German household structure (*(22)*, see supplementary material for detailed information on the household structure). Since the COVID-19 infectiousness typically declines within seven days after symptom onset *(21)*, individuals remain in the infectious state for nine days, before transitioning into the recovered state. Infectious individuals can infect susceptible individuals within a 2-meter distance. The likelihood of an infection event depends on the distance between an infectious and a susceptible individual and follows a half normal distribution (i.e., the smaller the distance, the more likely an infection event). Within households, we presume that an infectious family member always infects other members, as individuals within a household typically are within less than 2-meters distance for an extended period of time.

We also simulated the epidemic when CT solutions are applied. Overall, we simulated 46 different scenarios. These scenarios differ in the CT adoption level, the PDR for CT, and the probability of individuals stopping to use the CT app after a false positive quarantine. The initial CT adoption levels in our simulation scenarios are 20%, 40%, 60%, 80%, or 100% of the simulated population. Within each of these initial adoption levels, we simulated five different PDRs for CT apps. For four of the CT apps to register a contact, individuals need to be within a certain range for at least 15 seconds, specifically of 0.2, 1, 2, or 10-meters. For the fifth simulated CT app, individuals scan a sites-wide QR code provided at a location (i.e., workplace, school, supermarket) which changes once a day. In practice, each of these PDRs can be realized with sensors built-in in widely available smartphones. Table 1 provides an overview of possible realizations. We did not trace contacts within households, as we presume that household members communicate and quarantine in case another household member shows symptoms. To account for possible usage stops in case of false-positive quarantine, we conducted simulations with individuals stopping CT usage with a probability of either 25%, 50%, 75%, or 100% in case they were falsely quarantined. We simulated each scenario 30 times with different random seeds to obtain averaged results. Each simulation contained a society of 10,000 individuals per scenario and a time step of 100 ms. The code and random seeds we used for our simulation are available on GitHub, thus allowing other researchers to verify and build on our results *(35)*. Figure 1 exemplifies typical 2-day routines within our spatial simulation when an infection occurs on day one. Two example scenarios (short PDR and wide PDR) are illustrated. In both scenarios, the presence of a CT app potentially leads to wrong conclusions about individuals’ health status. In the short PDR scenario, the individuals D and H are potentially infectious but not tracked by CT (false negative) and in the wide PDR scenario, the individuals E and I are healthy but tracked by CT and subsequently quarantined (false positive).

**Fig. 1.**
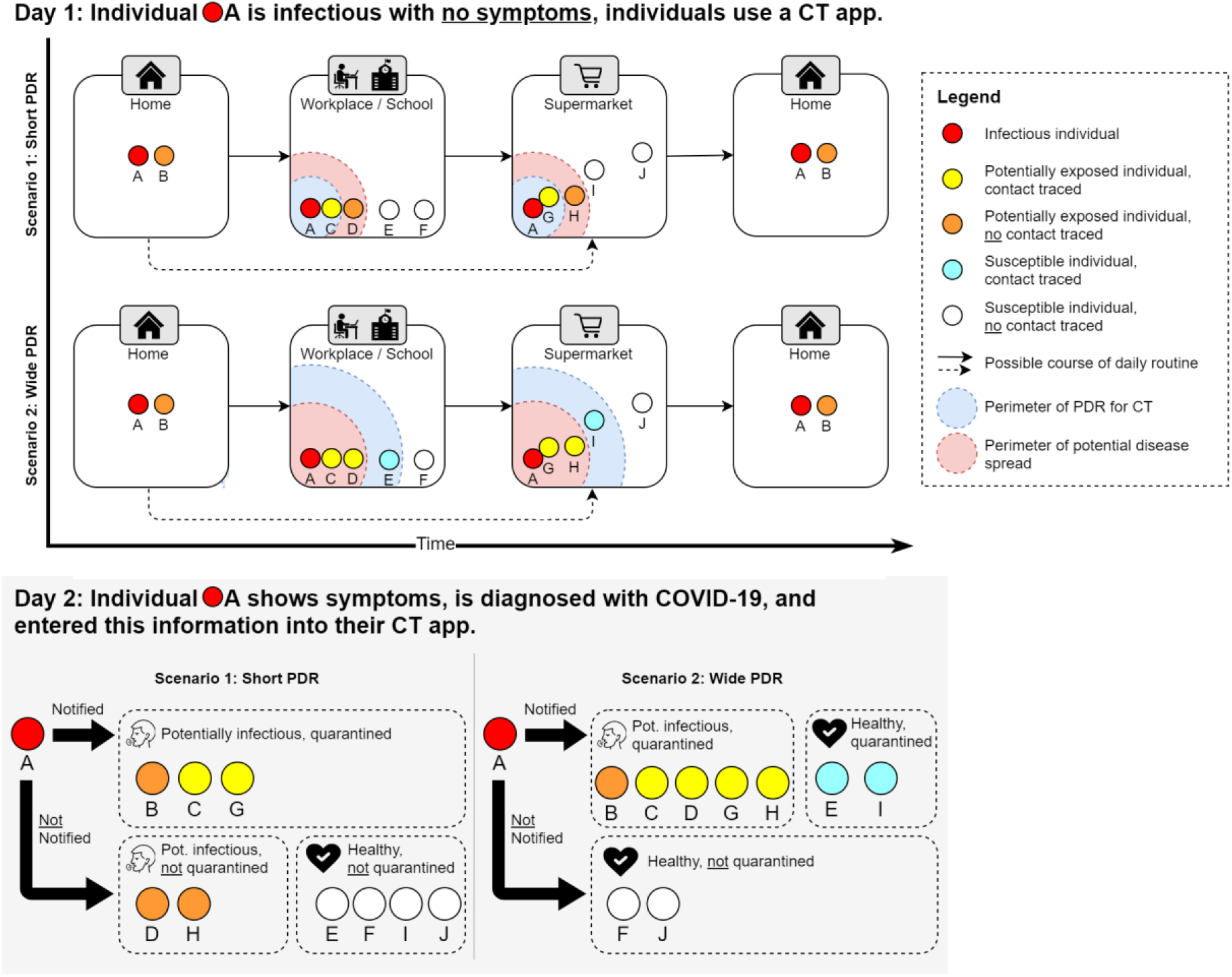
Illustration of the simulation. Illustration of CT apps with a small PDR, and a wide PDR. On day one, individual A is infectious but does not yet show symptoms. Therefore, individual A follows a daily routine and infects other individuals. Depending on the PDR of the CT apps, different contacts with other individuals are traced. At the end of day one, individual A develops symptoms and, thus, contacts are notified. On day two, individual A and the contacts go into quarantine, while non-contacts follow their daily routine.

### Effectiveness of contact tracing

A major goal of controlling the COVID-19 epidemic is to keep the maximum share of infectious people low, commonly referred to as ‘flattening the curve’. To analyze how different PDRs of different CT apps might influence the course of the COVID-19 epidemic, we simulated several CT solutions for each adoption level. The course of the average share of infectious individuals over time is illustrated in Figure 2. In this first simulation, we did not account for usage stops of individuals due to false positive quarantine.

**Fig. 2.**
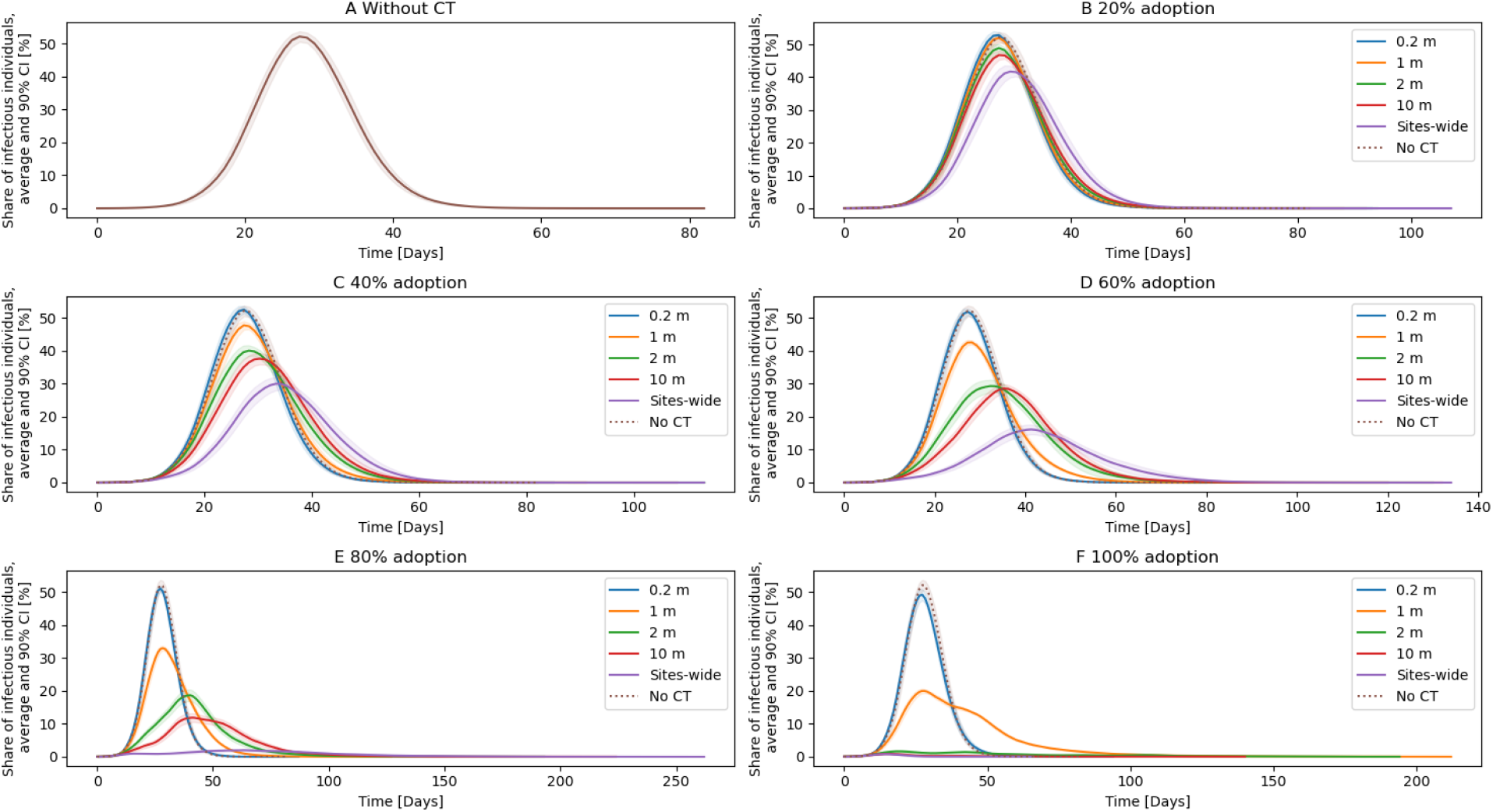
Average share of infectious individuals. For each curve, the mean from 30 simulations, and the 90% confidence intervals (displayed as colored shades behind each curve) are plotted. The characteristics of the individual curves depend highly on the PDR and the adoption rate.

The course of the simulated COVID-19 epidemic without any CT is illustrated in Figure 2A. The average maximum share of infectious individuals within the 30 simulations is 55.80%. With any CT solution adopted, this maximum decreases. In general, larger PDRs lead to a lower maximum share of infectious individuals than smaller PDRs. The reason for this is that large PDRs are more sensitive and cause more susceptible and infectious individuals to go into quarantine. The susceptible individuals then cannot get infected for the time of the quarantine, whereas the infectious individuals are prevented from infecting others (except for household members). Furthermore, the initial adoption rates of CT apps have a substantial impact on the course of the COVID-19 epidemic. Thereby, higher initial adoption rates lead to lower average maximum shares. For example, for CT based on a 10 m PDR, the average maximum is 50.20% at a 20% adoption rate (see Figure 2B) and 1.06% at a 100% adoption rate (see Figure 2F). The lowest maximum average of infectious individuals was 0.79% for the sites-wide PDR with a 100% adoption rate.

Another metric of interest is the duration of the epidemic, which is illustrated in Figure 3 A. Without CT, the average duration was 72.50 days. By flattening the curve, the adoption of CT apps generally led to a prolongation of the epidemic, with a higher adoption rate leading to an increase in the duration. The highest average duration was 164.47 days for a 1 m PDR at a 100% adoption rate. For CT based on PDRs of 2 m or larger, however, higher adoption did not necessarily lead to a prolonged epidemic. CT based on a sites-wide PDR, for example, had the shortest epidemic duration with an average of 41.73 days, followed by CT based on a 10 m PDR with an average of 56.20 days. Both values are shorter than the average duration of the epidemic without CT. The 2 m PDR CT solution adopted at 100% comes with an average epidemic duration of 95.77 days, which is more than the epidemic without CT, but less than the 2 m PDR adopted at 80 % (132.97 days).

**Fig. 3.**
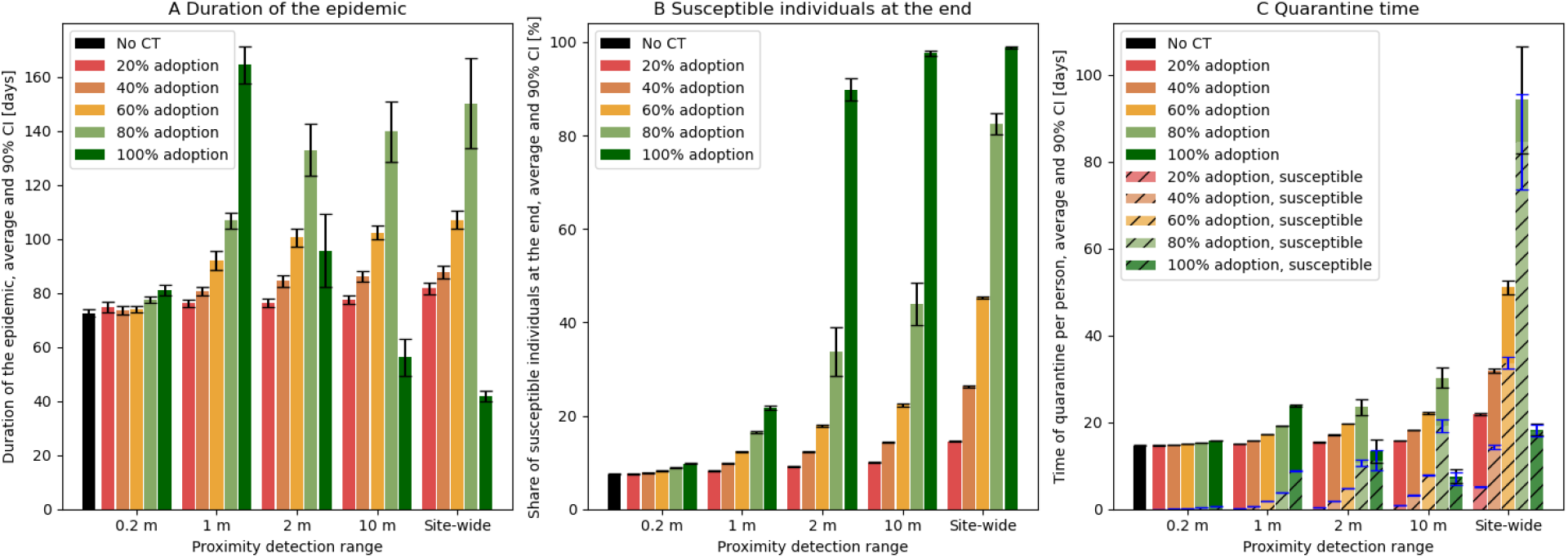
Quarantine time and efficiency measures. Each subfigure illustrates the mean values and 90% confidence intervals based on the used PDRs for CT and the adoption of CT averaged over 30 simulations. A shows the duration of the epidemic. For this, Figure 2 is not representative, as the curves are averaged and, therefore, take the value of the simulation with the longest duration. B shows the share of susceptible individuals at the end of the epidemic, C shows the average quarantine time per individual, and the average time individuals spend this quarantine time in a susceptible disease state.

We further analyzed the share of susceptible individuals at the end of the epidemic, illustrated in Figure 3B. A high share is preferable, since individuals that never get infected with the disease do not suffer from potential long-term consequences of an infection and will not succumb to the disease. Without any CT, almost all of the individuals in our simulation get infected over time, with the average share of susceptible individuals at the end of the simulation being 7.51%. Conversely, the higher the adoption rate of CT and the larger the PDR for CT, the higher the share of susceptible individuals at the end of the epidemic get. However, the effect of CT with small PDRs (i.e., 0.2 m, 1 m), is limited: With a 1 m PDR CT adopted at 100%, an average of 21.73% of the simulated population is susceptible at the end of the epidemic, whereas a rest (78.27%) has been infected over time. Contrary, a PDR of 2 m at a 100% adoption rate already reaches an average of 89.89% individuals susceptible at the end of the epidemic, and CT with the 10 m PDR reaches even an average of 97.68% susceptible individuals at the end of the epidemic. However, the value of the 10 m PDR CT is only 43.88% when adopted at 80%. With CT based on a sites-wide PDR, the value at 80% adoption rate is 82.57% and 98.82% at 100% adoption rate.

A major cost of CT is quarantine time, which can have significant negative effects, such as economic losses *(2, 11)*, strains on mental health *(12, 36)*, untreated medical conditions *(37)*, or increases in domestic violence *(38)*. Therefore, we analyzed the average quarantine time per person, which is illustrated in Figure 3C. Without CT, the average quarantine time per person was 14.69 days, due to quarantined infectious individuals, and their household members. In general, the average quarantine time per person increased with a higher CT adoption rate and larger PDRs. It reaches its maximum at 94.28 days for the sites-wide PDR with a 80% adoption rate. However, with adoption rates above 80%, CT based on a 2 m PDR, a 10 m PDR, or sites-wide PDR showed less quarantine time. At an adoption rate of 100%, the average quarantine time thereby is the lowest for CT based on a 10 m PDR with 7.55 days. CT based on a 2 m PDR resulted in 13.40 days, and CT based on sites-wide PDR resulted in 18.32 days of average quarantine time, which is slightly higher than the epidemic without any CT. Based on the average quarantine duration per person, we also analyzed what share of this time was spent in a susceptible state. In general, quarantine measures aim to target infectious individuals. It also seems reasonable to quarantine exposed individuals, as they may become infectious any time, and in practice it may be challenging to diagnose whether an individual is exposed but not infectious. However, one could consider a quarantine of susceptible individuals as false positive quarantine. The results of our analysis are illustrated in Figure 3C as hatched bars. Without CT, there are no susceptible individuals in quarantine, since only individuals with symptoms, which are also infectious, and their exposed household members go in quarantine. With CT, also susceptible individuals go into quarantine. For all PDRs, a higher adoption rate of CT leads to a higher share of susceptible individuals in quarantine. Furthermore, CT based on PDRs with a wider detection radius generally lead to a higher share of quarantine time spent in a susceptible state, as they register contacts at a relatively far away distance, where an actual infection event is unlikely to occur. The highest share of time spent in quarantine by susceptible individuals was 98.49% with CT based on the sites-wide PDR adopted at 100%. With a 2 m PDR based CT adopted at 100%, the share was still 85.15%.

### Contact tracing with a usage stop

After being in a quarantine and not feeling symptoms of sickness, an individual may perceive their quarantine as unnecessary. Eventually, quarantines perceived as unnecessary may, among other reasons (e.g., privacy concerns, general mistrust in a CT app or the government), lead to individuals stopping to use the CT app and a lower overall adoption rate of CT apps, thus impeding the effectiveness of CT. To account for this effect, we simulated usage stops after a false positive quarantine with different probabilities (25%, 50%, 75%, or 100%). In these simulations, we assumed an initial CT adoption rate of 60%. This is the recommended minimum adoption rate in other extant research *(2)*. At the same time, it appears as realistic in western societies, as a 2015 US-based survey revealed that 58.23% of mobile phone users had downloaded a health-related mobile app *(39)* and it is likely to assume that the smartphone adoption and health app usage has increased since then.

The results of these simulations are illustrated in Figure 4. Similar to the analysis conducted for Figure 2, we focus on the average share of infectious individuals over time. For CT with small PDRs (0.2 m, 1 m), the effect of this usage stop is almost non-existent. Without any usage stop, the average maximum was 54.71%, respectively 44.28%. With a 100% probability of a usage stop, these maxima are 54.72% and 44.26%. The small increase for the 0.2m PDR is likely due to a statistical variation and well within the 90% confidence interval. For CT based on a 2 m PDR with no usage stop, the average maximum is 32.49%. This value only slightly increases to up to 33.54% with a 100% probability usage stop. One reason for this observation may be that CT based on short PDR produces only very little false positives, and, thus, only very few individuals stopped using the CT app in our simulation. For CT based on larger PDRs, however, the usage stop has a severe effect on the average maximum share of infectious individuals. Without a usage stop, the values are 30.68% for a 10 m PDR and 19.48% for sites-wide PDR at 60% initial adoption. When studying a 25% probability of a usage stop, the 10 m PDR and the sites-wide PDR come with a 33.50% respectively 35.85% average maximum, thus being similarly effective than the 2 m PDR with 32.54%. This trend intensifies with an increasing probability of the usage stop, where especially the 10 m PDR and the sites-wide PDR become less effective. At a 100% probability, the CT based on sites-wide PDR comes with an average maximum share of 52.27% and is, thus, higher than CT based on a 2 m PDR with 33.54% and even higher than CT based on a 1 m PDR at 44.26%. CT based on a 10 m PDR lies in the middle of the latter two with 38.55%.

**Fig. 4.**
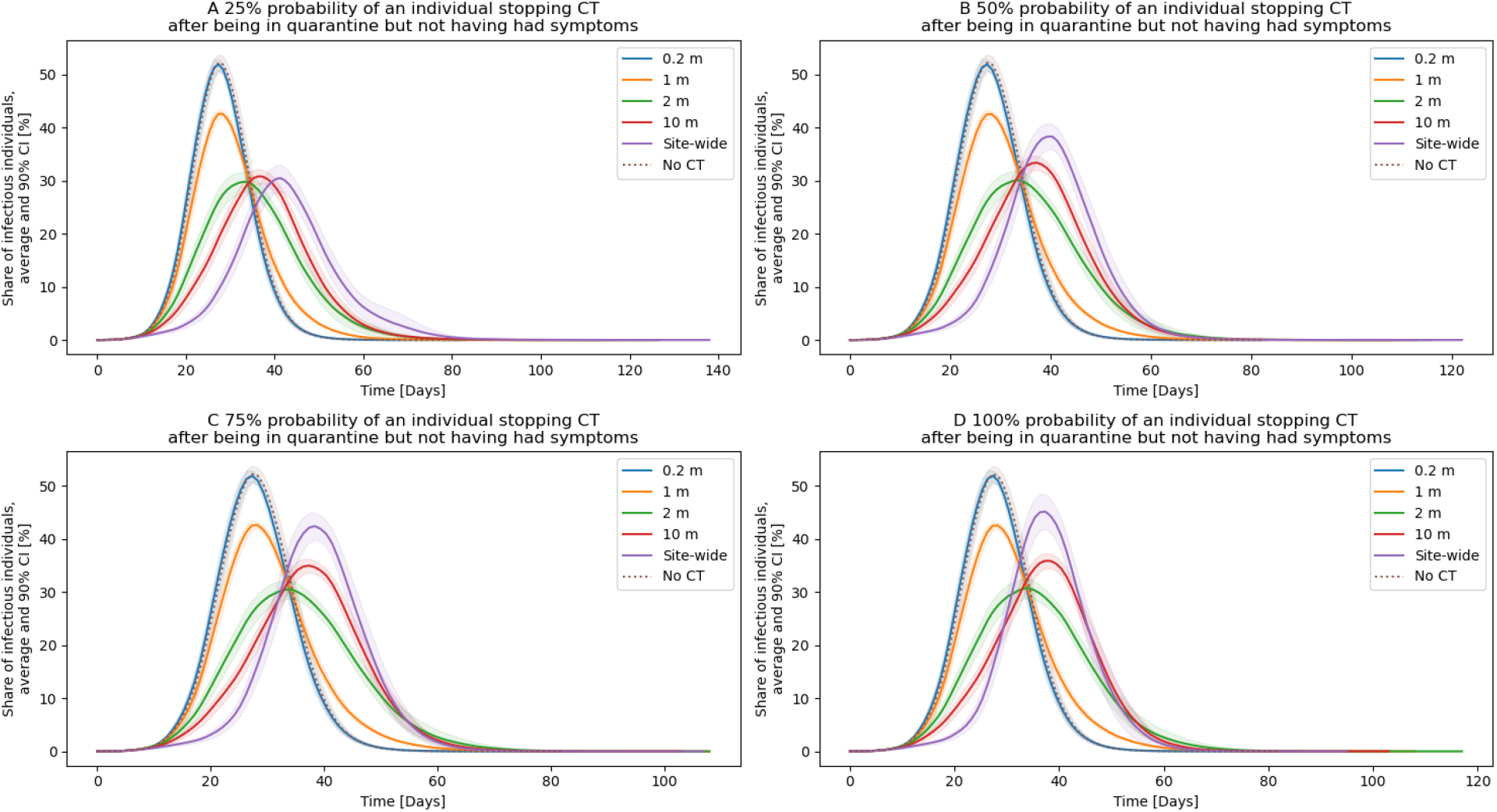
Usage stop of CT after being in a false positive quarantine. For each curve, the mean from 30 simulations, and the 90% confidence intervals are plotted. Thereby, the initial adoption rate of CT is 60%. The probability of stopping CT after a false positive quarantine varies in the subfigures.

## Discussion

With the simulation, we studied how various CT solutions based on different PDRs affect the course of an epidemic. Generally, the adoption of CT helped in all simulations to control the epidemic in the sense of decreasing the maximum share of infectious individuals, and increasing the share of susceptible individuals at the end of the epidemic. In doing so, introducing a CT app also helped to decrease the R_0_ value of the infectious disease within our simulation (see Table S2 in the supplementary materials for detailed information on R_0_ values). Our results are in line with previous research suggesting that high adoption rates of CT are beneficial *(33)*. While our results confirm prior results that a certain adoption rate is necessary to effectively stop an epidemic right from the beginning *(2)*, our research also suggests that there is no minimum threshold of CT to be effective, but that instead every participation can help to better control the epidemic and increase the effectiveness of CT itself.

The results of our spatial simulations show that each of the different PDRs yields its own advantages and disadvantages when it comes to fighting the COVID-19 epidemic. We summarize these advantages and disadvantages in Table 2. In general, a 0.2 m or 1 m detection range appear to only have limited effectiveness, illustrated, for example, by the share of susceptible individuals at the end of the pandemic, which is only 36.60% on average for the 1 m PDR even at 100% adoption rate. The 2 m, 10 m, and sites-wide PDR are all very effective in controlling the epidemic at 100% adoption rate. The sites-wide PDR is even highly effective at only 80% adoption rate. However, this effectiveness comes with several disadvantages such as very long quarantine duration, and high shares of susceptible individuals in quarantine (false positives). For 100% adoption rate of CT with a 10 m or sites-wide PDR, the average R_0_ value was actually below 1, meaning an infectious individual on average infects less than one other individual. Thus, both CT solutions enabled a declining epidemic right from the beginning. In all other simulation settings, the average R_0_ value always was above the critical threshold of 1, meaning not only CT, but also a growing herd immunity enabled an eventual decline of the epidemic. When simulating a usage stop with an initial adoption of 60%, the advantages and disadvantages of different PDRs for CT substantially changed. PDRs with a short detection range (0.2 m, 1 m) were unaffected, however, PDRs with a wide detection range (10 m, sites-wide) performed much worse. Under this scenario, our simulations demonstrated that the most effective CT was based on a 2 m PDR since the 10 m and the sites-wide PDR showed higher average maximum shares of infectious individuals and, thus, were less effective in ‘flattening the curve’.

**Table 2.** Simulated PDRs and their advantages and disadvantages for CT.

| PDR | Advantages | Disadvantages |
| --- | --- | --- |
| 0.2 m | • Lowest risk of detecting false positives | • Almost no positive impact on maximum share of infectious individuals, or share of susceptible individuals at the epidemic end |
| 1 m | • Noticeable decrease of the maximum share of infectious individuals at high adoption rates<br>• Relatively little quarantine time required<br>• Little share of susceptible individuals in quarantine | • Relatively low share of susceptible individuals at the end of the epidemic, even at 100% adoption rate<br>• Long epidemic duration at 100% adoption rate |
| 2 m | • Highly effective for epidemic control at 100% adoption rate<br>• Relatively little quarantine time required<br>• The most effective PDR for a 60% adoption rate with a usage stop | • Relatively high share of susceptible individuals in quarantine at 100% adoption<br>• Much less effective at lower adoption rates compared to sites-wide CT |
| 10 m | • Highly effective for epidemic control at 100% adoption rate<br>• The least amount of quarantine time required of all PDRs at 100% adoption rate | • Relatively high share of susceptible individuals in quarantine at an 80% adoption rate or higher<br>• Loses effectivity under a scenario of a usage stop because of false positive quarantines, even at just 25% probability |
| Sites-wide | • Highly effective for epidemic control, when considering the maximum share of infectious individuals and the share of susceptible individuals at the end, even at 80% adoption rate<br>• Out of all PDRs, the lowest epidemic | • Highest share of quarantine time by susceptible individuals of more than 50% at only 60% adoption rate or higher<br>• Relatively long quarantine times at an adoption rate of 60% or 80%<br>• Strongly loses its effectivity at 60% initial adoption |
|  | duration at 100% adoption rate | and a usage stop effect |

Our results imply that there is no silver bullet CT technology. Instead, choosing the right CT technology always means finding a compromise between various factors such as effectiveness in controlling the epidemic, false positive quarantine, share of susceptible people, and privacy concerns. In this regard, our work can help decision makers to assess the advantages and disadvantages of the different technologies and, thus, make more informed decisions. Different countries with different cultures have shown widely varying acceptance rates of COVID-19 measures in general (e.g., lockdown, mask requirements), and varying adoption rates of CT apps in particular. Factors influencing the adoption rates may include privacy concerns, the perception of the CT app’s effectiveness, and the perceived quarantine time required to comply with CT apps. Furthermore, some CT technologies may be easy to enforce (e.g., scanning a QR code before entering places of everyday life), which could increase the adoption rate and effectivity of a CT app, compared to other CT technologies. In some countries, CT based on wide PDRs (i.e., 10 m, sites-wide) may have higher adoption rates than CT based on narrower PDRs due to their higher effectiveness, high acceptance of false positive quarantine, or their absence of causing highly precise movement profiles. Bluetooth Class 2, GPS, wider versions of Bluetooth Low Energy, or sites-wide QR codes may be appropriate technologies for CT apps in such countries (see Table 1). In other countries, however, there may be a high chance of a usage stop after a false positive quarantine. In such cases, our results support the adoption of PDRs that have at least a range of a typical disease spread, but not much more. Bluetooth Low Energy may be an appropriate technology for CT apps in such countries. The results of our simulations also show that technologies with very short PDRs such as NFC are not suitable for CT in the case of the COVID-19 epidemic since they resulted in almost no positive impact on the maximum share of infectious individuals, or share of susceptible individuals at the end.

With our work, we follow a recent call for research on the technical design of CT apps *(10)* and open multiple opportunities for future research. However, our work is limited by some factors as the real world is always more complex than a simulation. For example, our simulation especially accounted for droplet infection, where CT can be highly effective. Recent research suggest that aerosol transmission or fomite transmission are another plausible mode of SARS-CoV-2 transmission *(40)*. As these modes of transmission do not directly depend on the physical proximity between individuals, CT may likely be less effective for epidemic control in such scenarios. Furthermore, contact patterns in the real world are likely to be more complex than in our simulations. Future research could, therefore, aim to develop more accurate measures for contact patterns and incorporate these into simulations; we made our simulation source code publicly available for such purposes. In general, the research on SARS-CoV-2 transmission is still ongoing. For our simulations, we required a probability distribution which tells us the likelihood of an infection, depending on the distance between the infectious and a susceptible individual. Our most likely choice was a half normal distribution which we parametrized such that we obtained an *R*_*0*_ value realistic for COVID-19. While we believe our choice is reasonable, future research could aim to further quantify this probability distribution with real-world evidence. However, such viral experiments may be challenging to conduct. Finally, recent research suggests that already recovered individuals can get infected with SARS-CoV-2 for a second time. Therefore, another interesting extension of our work would be the usage of a SEIRS model instead of a SEIR model, where individuals can transition to the susceptible state from the recovered state.

Extant research has demonstrated that the economic and societal consequences of large lockdowns with many quarantined people are highly severe. In this regard, we demonstrate that choosing the right CT technology with a suitable PDR can play a key role in determining whether large lockdowns become necessary or not. On the one hand, short PDRs are limited in their effectiveness of fighting the epidemic since they potentially miss to track important contacts. On the other hand, wide PDRs should also be selected with care since they lead to high shares of false positive quarantine. Our results suggest, that for many scenarios, the most promising CT solutions are based on a PDR that roughly corresponds to the infection range. However, this finding is dependent on various different factors such as initial adoption, and potential usage stops. Consequently, we recommend further exploration of reasons for such usage stops and means to ensure extensive continuous use of CT apps.

## Data Availability

The paper (with the supplementary material) describes the research method, and the source code for the simulation is linked to GitHub.

https://github.com/kpandl/COVID-19-contact-tracing-simulation

## Acknowledgments

The authors acknowledge support by the state of Baden-Württemberg through the high performance computing cluster bwHPC.

## Funding

This research is partially funded by the German Research Foundation (DFG). Grant number: DFG SU 717/15-1;

## Author contributions

Conceptualization: K.P., S.T., Data Curation: K.P., Formal analysis: K.P., M.S., Funding acquisition: S.T., A.S., Investigation: K.P., Methodology: K.P., S.T., M.S., Project administration: K.P., Resources: A.S., Software: K.P., Supervision: S.T., Visualization: K.P., S.T., M.S., Writing, original draft: all authors, Writing, review, and editing: all authors.

## Competing interests

None declared.

## Data and materials availability

All data are available in the manuscript or the supplementary materials. The software code used for our analyses is publicly available *(35)*. This work is licensed under a Creative Commons Attribution 4.0 International (CC BY 4.0) license, which permits unrestricted use, distribution, and reproduction in any medium, provided the original work is properly cited. To view a copy of this license, visit https://creativecommons.org/licenses/by/4.0/. This license does not apply to figures/ photos/artwork or other content included in the article that is credited to a third party; obtain authorization from the rights holder before using such material.

## Supplementary Materials for

### Materials and methods

#### General simulation model

Our simulation model consists of a population of 10,000 individuals living in households. The household size distribution follows the German household size distribution *(22)* and is illustrated in Table S1. We assume that individuals return to their household every day.

**Table S1.** Household sizes and their share of the total number of households within the simulation.

| Household size [number of individuals] | Share of total number of households [%] |
| --- | --- |
| 1 | 42.3 |
| 2 | 33.2 |
| 3 | 11.9 |
| 4 | 9.1 |
| 5 | 3.5 |

Following prior work on the spread of infectious diseases *(16)*, individuals can meet at four different types of places. In our simulation, these places are workplaces, schools, supermarkets, and households. Similar to the German society, we assumed that 70.1% of the population regularly visit a workplace or school (including universities and kindergartens). This estimated share is based on ca. 44.7 million individuals working *(41)*, ca. 8.3 million pupils in school *(42)*, ca. 2.8 million university students *(43)*, and 2.4 million kindergarteners *(44)*, out of a German population of 83 million individuals. In our simulation, these workplaces or schools have a quadratic shape (as illustrated in Figure S1 B) with a row capacity between 2 and 8, thus, the total capacity of the specific location is between 4 and 64. When visiting a workplace or school, an individual randomly selects one seat, which ensures a mixing of different contacts in our simulation. The distance between each row and column for a workplace or school are randomly sampled between 1.5 and 2-meters. Thereby, individuals can move along aisles in order to go to their seats with a constant speed of 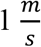. Individuals arrive between 8 AM and 8:30 AM and stay there for 8 hours.

**Fig. S1.**
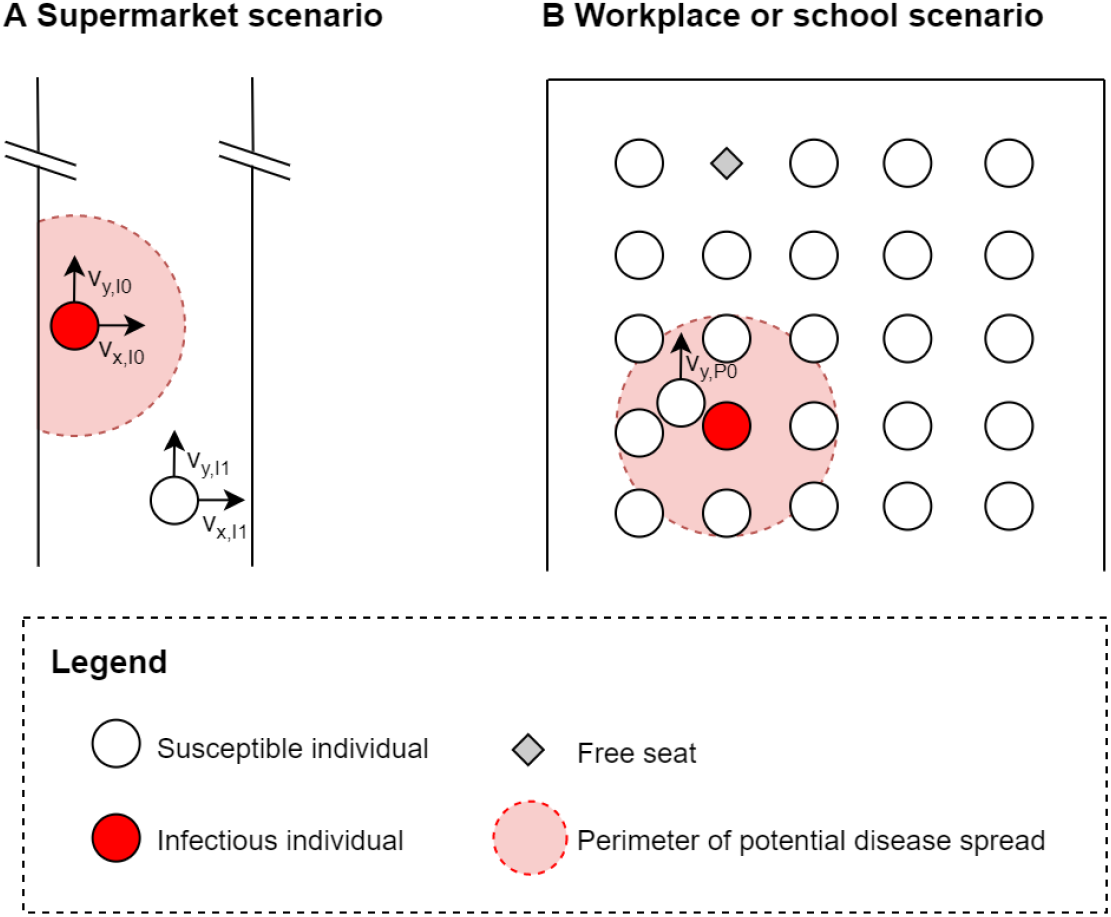
Illustration of the spatial simulation locations. Besides households, infectious individuals can infect other individuals at supermarkets, workplaces, and schools. This figure shows individuals at these places, some of the shown individuals are moving, illustrated by a speed component in x and y direction. Specifically, A shows a supermarket scenario, where individuals move along an aisle. B shows a workplace or school scenario in quadratic shape, where an individual moves to a randomly assigned seat when arriving, and may get infected while doing so.

If an individual is not at their workplace or school, they can visit a supermarket once a day with a daily probability of 20%. These are open Monday through Saturday from 8 AM till 8 PM. For the 10,000 individuals in the population, there are three supermarkets, resulting in a comparable distribution of supermarkets per capita to Germany *(45)*. A supermarket is simulated as one long aisle, as illustrated in Figure S1 A. Individuals can go in both directions, forward, and backward, which increases the mixture of contacts. To do so, each individual has a constant base speed in y direction, which is determined such that the planned duration of the visit is a random number between 15 and 45 minutes. To allow individuals walking backwards, an individual walks with an acceleration in y direction which takes a sinusoidal form. The period duration is three minutes, the amplitude is determined such that the maximum additional speed gain from this sinusoidal function is the difference to the maximum speed of 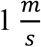. However, this maximum additional speed gain is divided by a random integer between 1 and 10. As a result, some individuals occasionally walk backward (meaning they walk with a negative speed), whereas others walk almost with a straight speed. The x acceleration is determined randomly, but capped such that the maximum total speed (consisting of a speed in x direction and a speed in y direction) is 1 m/s. Furthermore, the individuals always stay within the physical boundaries.

In general, the next position of a person as a vector is determined by its current speed 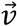, its acceleration 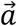, and the simulation time step *Δt*:

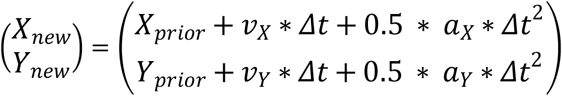

The speed vector of the person after the next time step is defined as follows:

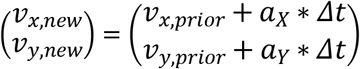

The time step in our simulation *Δt* is 100 ms, as this allows us to simulate walking speeds in the order of about 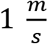 and measure distances in the order of decimeters at the same time.

#### Epidemic simulation

Another essential part of our simulation model is the infectious disease. We aim to model this infectious disease realistic to COVID-19 with a SEIR model, where individuals are either in a susceptible, an exposed, an infectious, or a recovered state. This flow of states is also illustrated in Figure S2.

**Fig. S2.**
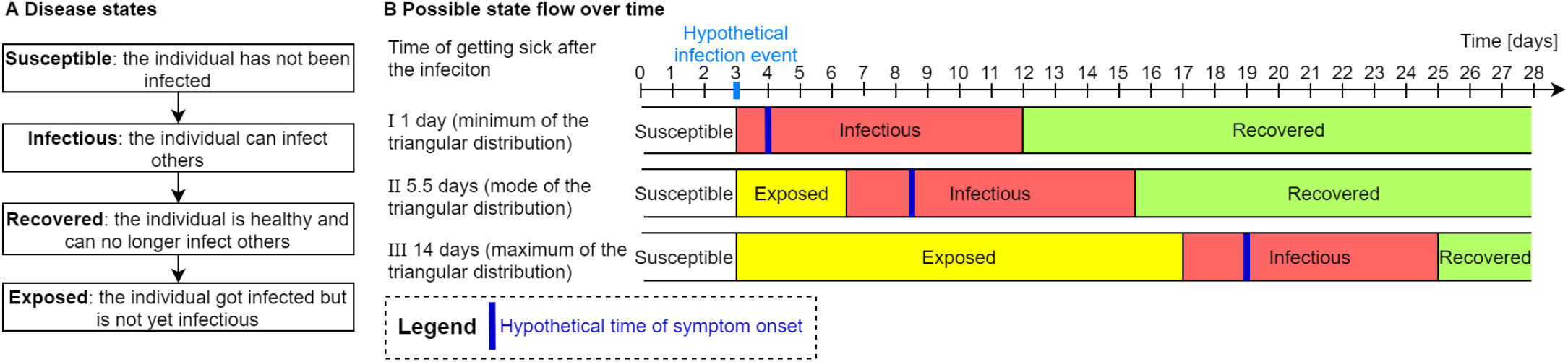
A shows the disease states and flow of the SEIR model. B shows different cases (minimum, mode, and maximum of the triangular distribution) on a time line.

At the start of the simulation, ten individuals of the population are exposed and the rest of the population is susceptible. The transition of a susceptible state to an exposed state occurs when an individual gets infected. Afterwards, it takes a certain duration until the individual develops symptoms. In accordance with extant knowledge on COVID-19 *(20)*, we model this duration as a random number sampled from a triangular distribution with a minimum of 1 day, a mode of 5.5 days, and a maximum of 14 days. Similarly, two days before an individual develops symptoms, its disease status transits from the exposed state to the infectious state. However, the duration in the exposed state cannot be negative. The duration in the infectious state is 9 days, effectively meaning 2 days before the symptoms and 7 days after the symptoms are noticed *(21)*. We implemented the spatial simulation for the workplaces, schools, and supermarkets, but did not implement it for the households. Instead, we assume that household members typically have a close contact between each other, therefore, in the simulation model an individual infects other household members once they are infectious. Furthermore, an individual and their household members go into quarantine the day after an individual showed symptoms.

A further important set of parameters determines the probability of an infection event. For such an infection event, an infectious and a susceptible individual need to be within a certain distance and the closer they are, the more likely the infection event occurs. Specifically, an infection event can occur only if the two individuals are in proximity of 2 m or closer. In our simulation, each infectious individual is assigned a fixed random value *σ* from a triangular distribution with a minimum of 0, a mode of 0.5 m, and a maximum of 1 m. This value accounts for a varying infectiousness between individuals which is observed for COVID-19 *(46)*. The higher it is, the more infectious an individual is, given it is in the infectious state of the disease. Once a susceptible individual is in proximity of an infectious individual, a random number from a half normal distribution is drawn. This half normal distribution has a standard deviation of *σ* from the infectious individual. An infection event can then occur if the distance between the two individuals is smaller than this drawn random number. This mechanism alone, however, would lead many more infections if the simulation time step *Δt* is small (it is 100 ms in our simulations). Therefore, another condition has to be met: a random event of 5% probability needs to be drawn to be true. The first condition ensures that an infection event is more likely, the closer two individuals are. A half normal distribution was chosen instead of a normal distribution because the distance between individuals is always a non-negative value. The second condition is necessary in order to adapt the likelihood of an infection to the simulation time step. Based on 30 simulations without any CT, this setting results in in an average base reproduction number of R_0_ = 2.799, a median of R_0_ = 2.792, and a 90% confidence interval between 2.787 and 2.811. The median value in our simulation was close to the median value of 2.79 that was identified for the SARS-CoV-2 pandemic by extant research *(17)*.

#### Measurement of the base reproduction rate

As discussed above, we set a few parameters such that we obtain a reasonable value for the base reproduction number R_0_. This affects parameters that define the likelihood of an infection, given an infectious individual is in proximity of a susceptible individual. This section describes how we calculated the base reproduction rate *R*_0_ out of our simulation results.

In a simplified SEIR model *(18, 47)* with a natural death rate of µ = 0, the base reproduction rate is given by 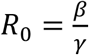. Thereby, *γ* is the reciprocal of the infectious period and *β* is the effective contact rate. The SEIR model is described by a system of differential equations, which themselves describe the change of four quantities. These four quantities are the number of susceptible individuals at a given time *S(t)*, as well as the number of exposed individuals *E(t)*, the number of infectious individuals *I(t)*, and the number of recovered individuals at a given time *R(t)*. The system of differential equations is as follows *(N* is the size of the population, the average incubation period is *a*^−1^):

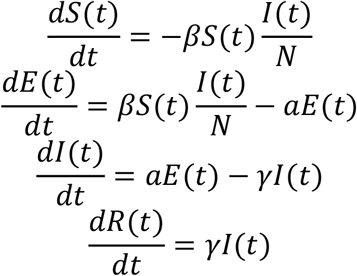

By dividing 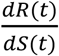, we obtain 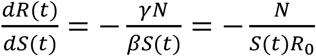. We then integrate from the start of the simulation *(t* = 0) to the end of the simulation *(t* = ∞) and obtain 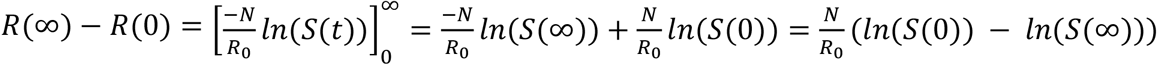. With *R*(∞) = *N* − *S*(∞) and *R*(0) = 0, we obtain 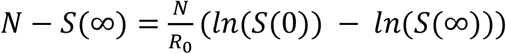, and ultimately, we obtain 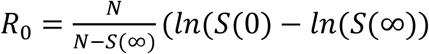. Thus, we can calculate *R*_0_ by the number of individuals in our simulated population *N* = 10,000, the number of initially susceptible individuals *S*(0) = 9,990, and the number of susceptible individuals at the end of the simulation *S*(∞).

#### Simulation process

We simulated on a high-performance computing cluster with nodes consisting of an Intel Xeon Gold 6230 CPU at 2.1 GHz frequency. In total, we simulated 46 different types of scenarios (composed of different CT adoption rates, different PDRs, and varying probabilities of a usage stop) Each scenario type was simulated 30 times (with different random seeds), in order to obtain smoothed means and confidence intervals. Each simulation ran on one CPU core, therefore, we computed with 1,380 CPU cores in total. Depending on the simulation parameters and random influences, the simulation time varied, most of the simulations finished within 1 and 2 days, the longest one took ca. 7.8 days. In order to decrease the required simulation time, one could increase the simulation time step *Δt* (e.g., to 1 s instead of 100 ms), and one could reduce the size of population. However, this can lead to less accurate results, and a higher variance; this is the reason we set the simulation time step to 100 ms. The source code to simulate and visualize the results is available at *(35)*.

### Simulation results

Table S2 shows the main results of our study with the mean value and 90 % confidence intervals based on 30 simulations. Most of these results are also visualized in Figure 2, 3, and 4 within the main article.

**Table S2.** Results of the different simulations, and 90 % confidence intervals.

| Initial adoption rate [%] | Usage stop probability [%] | PDR | Maximum share of Infectious individuals [%] | Time till maximum share of infectious individuals | Duration of the pandemic [days] | Average time of quarantine per person [days] | Share of susceptible quarantine time [%] | Share of susceptible people at the end [%] | Overall R0 value □ |
| --- | --- | --- | --- | --- | --- | --- | --- | --- | --- |
| 0 | 0 | No CT | 55.796 ± 0.145 | 27.367 ± 64.173 | 72.500 ± 1.415 | 14.690 ± 0.025 | 0.000 ± 0.000 | 7.511 ± 0.103 | 2.799 ± 0.012 |
| 20 | 0 | 0.2 m | 55.659 ± 0.232 | 26.633 ± 55.641 | 74.800 ± 1.798 | 14.722 ± 0.028 | 0.084 ± 0.006 | 7.585 ± 0.095 | 2.790 ± 0.011 |
| 20 | 0 | 1 m | 54.545 ± 0.229 | 26.900 ± 57.692 | 76.200 ± 1.524 | 15.019 ± 0.028 | 1.046 ± 0.032 | 8.165 ± 0.084 | 2.728 ± 0.009 |
| 20 | 0 | 2 m | 52.069 ± 0.267 | 27.167 ± 66.745 | 76.533 ± 1.547 | 15.432 ± 0.031 | 3.209 ± 0.067 | 9.130 ± 0.109 | 2.634 ± 0.010 |
| 20 | 0 | 10 m | 50.169 ± 0.210 | 27.533 ± 73.458 | 77.467 ± 1.604 | 15.818 ± 0.032 | 5.722 ± 0.099 | 9.979 ± 0.118 | 2.560 ± 0.010 |
| 20 | 0 | Sites-wide | 46.565 ± 0.278 | 29.667 ± 90.212 | 81.667 ± 2.232 | 21.899 ± 0.199 | 23.636 ± 0.499 | 14.500 ± 0.155 | 2.258 ± 0.008 |
| 40 | 0 | 0.2 m | 55.301 ± 0.235 | 26.733 ± 60.532 | 73.433 ± 1.556 | 14.829 ± 0.028 | 0.366 ± 0.018 | 7.793 ± 0.090 | 2.767 ± 0.010 |
| 40 | 0 | 1 m | 50.846 ± 0.277 | 27.500 ± 62.879 | 80.567 ± 1.501 | 15.837 ± 0.030 | 4.478 ± 0.086 | 9.823 ± 0.122 | 2.573 ± 0.011 |
| 40 | 0 | 2 m | 43.738 ± 0.303 | 28.333 ± 83.640 | 84.500 ± 2.153 | 17.111 ± 0.035 | 11.499 ± 0.151 | 12.227 ± 0.132 | 2.394 ± 0.009 |
| 40 | 0 | 10 m | 40.453 ± 0.310 | 30.233 ± 94.238 | 86.167 ± 2.085 | 18.173 ± 0.045 | 17.736 ± 0.227 | 14.291 ± 0.154 | 2.269 ± 0.008 |
| 40 | 0 | Sites-wide | 34.066 ± 0.333 | 33.767 ± 128.989 | 87.767 ± 2.356 | 31.920 ± 0.584 | 44.787 ± 0.734 | 26.254 ± 0.255 | 1.813 ± 0.007 |
| 60 | 0 | 0.2 m | 54.714 ± 0.285 | 26.867 ± 57.892 | 74.133 ± 1.208 | 15.030 ± 0.031 | 0.979 ± 0.037 | 8.187 ± 0.122 | 2.726 ± 0.013 |
| 60 | 0 | 1 m | 44.276 ± 0.317 | 27.767 ± 53.548 | 92.100 ± 3.621 | 17.206 ± 0.039 | 10.676 ± 0.123 | 12.306 ± 0.151 | 2.389 ± 0.010 |
| 60 | 0 | 2 m | 32.488 ± 0.277 | 32.267 ± 145.062 | 100.533 ± 3.211 | 19.700 ± 0.070 | 24.091 ± 0.210 | 17.828 ± 0.194 | 2.098 ± 0.008 |

| Initial adoption rate [%] | Usage stop probability [%] | PDR | Maximum share of Infectious individuals [%] | Time till maximum share of infectious individuals | Duration of the pandemic [days] | Average time of quarantine per person [days] | Share of susceptible quarantine time [%] | Share of susceptible people at the end [%] | Overall R0 value [] |
| --- | --- | --- | --- | --- | --- | --- | --- | --- | --- |
| 60 | 0 | 10 m | 30.678 ± 0.386 | 35.800 ± 116.680 | 102.300 ± 2.536 | 22.138 ± 0.154 | 35.126 ± 0.497 | 22.292 ± 0.276 | 1.931 ± 0.009 |
| 60 | 0 | Sites-wide | 19.478 ± 0.283 | 42.567 ± 216.113 | 107.033 ± 3.411 | 51.062 ± 1.541 | 65.887 ± 0.754 | 45.340 ± 0.299 | 1.445 ± 0.004 |
| 80 | 0 | 0.2 m | 53.281 ± 0.280 | 26.900 ± 53.360 | 77.600 ± 1.269 | 15.380 ± 0.036 | 2.146 ± 0.055 | 8.843 ± 0.105 | 2.661 ± 0.010 |
| 80 | 0 | 1 m | 34.515 ± 0.328 | 27.933 ± 67.127 | 106.900 ± 2.958 | 19.229 ± 0.066 | 20.137 ± 0.191 | 16.458 ± 0.261 | 2.160 ± 0.012 |
| 80 | 0 | 2 m | 19.865 ± 1.624 | 38.667 ± 235.762 | 132.967 ± 9.737 | 23.486 ± 1.825 | 47.843 ± 3.045 | 33.764 ± 5.273 | 1.683 ± 0.066 |
| 80 | 0 | 10 m | 14.144 ± 1.179 | 41.867 ± 323.817 | 139.800 ± 11.154 | 30.284 ± 2.369 | 65.305 ± 2.397 | 43.879 ± 4.518 | 1.483 ± 0.053 |
| 80 | 0 | Sites-wide | 2.810 ± 0.421 | 62.167 ± 769.469 | 150.200 ± 16.753 | 94.284 ± 12.250 | 90.813 ± 1.178 | 82.570 ± 2.286 | 1.070 ± 0.033 |
| 100 | 0 | 0.2 m | 52.227 ± 0.292 | 26.933 ± 68.895 | 81.033 ± 1.803 | 15.793 ± 0.039 | 3.627 ± 0.076 | 9.744 ± 0.112 | 2.580 ± 0.010 |
| 100 | 0 | 1 m | 21.385 ± 0.396 | 27.667 ± 93.485 | 164.467 ± 6.733 | 23.831 ± 0.162 | 36.597 ± 0.341 | 21.732 ± 0.403 | 1.951 ± 0.014 |
| 100 | 0 | 2 m | 2.391 ± 0.386 | 37.900 ± 738.780 | 95.767 ± 13.384 | 13.404 ± 2.720 | 85.156 ± 0.757 | 89.891 ± 2.340 | 1.026 ± 0.025 |
| 100 | 0 | 10 m | 1.059 ± 0.190 | 20.933 ± 516.769 | 56.200 ± 6.819 | 7.553 ± 1.616 | 93.846 ± 0.369 | 97.683 ± 0.574 | 0.928 ± 0.022 |
| 100 | 0 | Sites-wide | 0.791 ± 0.149 | 13.900 ± 43.419 | 41.733 ± 1.896 | 18.326 ± 1.421 | 98.489 ± 0.242 | 98.823 ± 0.224 | 0.885 ± 0.020 |
| 60 | 25 | 0.2 m | 54.714 ± 0.285 | 26.867 ± 57.892 | 74.633 ± 1.264 | 15.031 ± 0.031 | 0.979 ± 0.038 | 8.156 ± 0.128 | 2.729 ± 0.013 |
| 60 | 25 | 1 m | 44.342 ± 0.302 | 27.800 ± 55.052 | 91.167 ± 2.817 | 17.141 ± 0.040 | 10.308 ± 0.117 | 11.901 ± 0.148 | 2.416 ± 0.010 |
| 60 | 25 | 2 m | 32.784 ± 0.319 | 32.733 ± 145.475 | 100.033 ± 2.699 | 19.507 ± 0.053 | 21.633 ± 0.177 | 14.079 ± 0.165 | 2.281 ± 0.009 |
| 60 | 25 | 10 m | 33.499 ± 0.390 | 36.767 ± 108.477 | 103.767 ± 2.251 | 21.227 ± 0.074 | 28.903 ± 0.277 | 15.151 ± 0.190 | 2.224 ± 0.010 |
| 60 | 25 | Sites-wide | 35.853 ± 0.853 | 41.500 ± 177.638 | 111.567 ± 3.488 | 33.797 ± 0.260 | 42.756 ± 0.651 | 16.801 ± 0.341 | 2.145 ± 0.016 |
| 60 | 50 | 0.2 m | 54.714 ± 0.285 | 26.867 ± 57.892 | 75.233 ± 1.313 | 15.034 ± 0.032 | 0.977 ± 0.038 | 8.145 ± 0.134 | 2.730 ± 0.014 |
| 60 | 50 | 1 m | 44.245 ± 0.309 | 27.700 ± 51.526 | 89.867 ± 2.140 | 17.088 ± 0.045 | 10.126 ± 0.111 | 11.687 ± 0.161 | 2.431 ± 0.011 |
| 60 | 50 | 2 m | 33.039 ± 0.342 | 32.867 ± 141.664 | 95.767 ± 2.093 | 19.269 ± 0.047 | 20.228 ± 0.138 | 11.959 ± 0.149 | 2.412 ± 0.010 |
| 60 | 50 | 10 m | 35.766 ± 0.523 | 37.100 ± 97.273 | 98.900 ± 2.402 | 20.572 ± 0.048 | 25.934 ± 0.229 | 11.857 ± 0.120 | 2.418 ± 0.008 |
| 60 | 50 | Sites-wide | 44.312 ± 0.790 | 39.400 ± 116.474 | 96.800 ± 2.635 | 26.408 ± 0.086 | 37.088 ± 0.431 | 11.010 ± 0.192 | 2.480 ± 0.015 |
| 60 | 75 | 0.2 m | 54.717 ± 0.286 | 26.867 ± 57.892 | 75.100 ± 1.496 | 15.029 ± 0.033 | 0.976 ± 0.038 | 8.169 ± 0.129 | 2.728 ± 0.013 |
| 60 | 75 | 1 m | 44.278 ± 0.312 | 27.800 ± 52.822 | 90.167 ± 2.324 | 17.052 ± 0.052 | 9.931 ± 0.099 | 11.485 ± 0.204 | 2.445 ± 0.014 |
| 60 | 75 | 2 m | 33.203 ± 0.367 | 33.300 ± 144.067 | 96.467 ± 1.749 | 19.122 ± 0.039 | 19.418 ± 0.147 | 10.375 ± 0.118 | 2.528 ± 0.009 |
| 60 | 75 | 10 m | 37.526 ± 0.614 | 37.033 ± 93.211 | 91.567 ± 1.598 | 20.130 ± 0.037 | 24.541 ± 0.185 | 10.147 ± 0.139 | 2.546 ± 0.011 |
| 60 | 75 | Sites-wide | 48.984 ± 0.609 | 38.033 ± 111.968 | 87.600 ± 1.878 | 23.205 ± 0.057 | 33.234 ± 0.289 | 8.807 ± 0.098 | 2.664 ± 0.009 |
| 60 | 100 | 0.2 m | 54.718 ± 0.286 | 26.867 ± 57.892 | 74.600 ± 1.427 | 15.026 ± 0.032 | 0.975 ± 0.038 | 8.192 ± 0.126 | 2.725 ± 0.013 |
| 60 | 100 | 1 m | 44.259 ± 0.324 | 27.700 ± 54.365 | 92.200 ± 2.039 | 17.044 ± 0.037 | 9.782 ± 0.107 | 11.175 ± 0.164 | 2.467 ± 0.012 |
| 60 | 100 | 2 m | 33.543 ± 0.439 | 33.033 ± 148.047 | 93.100 ± 2.244 | 18.970 ± 0.037 | 19.014 ± 0.119 | 9.903 ± 0.129 | 2.566 ± 0.011 |
| 60 | 100 | 10 m | 38.548 ± 0.731 | 37.567 ± 90.660 | 90.233 ± 1.596 | 19.875 ± 0.033 | 23.780 ± 0.177 | 9.044 ± 0.106 | 2.642 ± 0.010 |
| 60 | 100 | Sites-wide | 52.271 ± 0.422 | 37.233 ± 113.356 | 82.667 ± 1.407 | 21.641 ± 0.041 | 30.365 ± 0.190 | 7.834 ± 0.077 | 2.763 ± 0.009 |

## Notes

### Competing Interest Statement

The authors have declared no competing interest.

